# Cross-cultural adaptation of the Arabic version of the Waterloo Footedness Questionnaire-Revised: WFQ-Ar

**DOI:** 10.1101/2023.01.04.23284187

**Authors:** Mishal M. Aldaihan

## Abstract

**Background and purpose:** Evaluating footedness is of great importance to clinical and behavioural research. The purpose of the current study is to translate the Waterloo Footedness Questionnaire-Revised (WFQ-R) to the Arabic language and investigate its psychometric properties.

**Methods:** Two independent forward translations were performed by two native Arabic speakers; and then synthesized into one version. The synthesized version was back translated into English by two independent bilingual translators. An expert committee was formed to review the translation and adaptation process. A final Arabic version of the WFQ-R was obtained. In order to test the internal consistency, reliability, and validity of the Arabic WFQ-R, adult Arabic speakers were recruited to participate in this study.

**Results:** For the cross-cultural adaptation, only one item was changed in order to express its conceptual meaning. Analysis showed an absence of floor and ceiling effect for the Arabic WFQ-R. Results of construct validity showed that all items of the translated WFQ-R have one dimension. For internal consistency of the Arabic WFQ-R, Cronbach’s alpha was excellent (0.93). ICC values showed excellent test-retest reliability (0.94). The Bland-Altman plot showed acceptable agreement between test and retest scores.

**Conclusion:** The Arabic WFQ-R is valid, reliable and ready for use among the Arabic speaking population for determining footedness.

## INTRODUCTION

Foot preferences or footedness is one of the behavioural predictors of cerebral functional laterality [1]. Laterality refers to the functional asymmetric use or preferred use of paired limbs or sensations. Four behavioural predictors are commonly used to determine functional laterality: footedness, handedness, earedness and eyedness [1]. Neuropsychology and motor behaviour studies indicates that laterality preferences in footedness, handedness, earedness and eyedness are interrelated [1-11]. Further, among the interrelation between the aforementioned laterality preferences, footedness and handedness show the strongest association [1-11]. Previous research indicates that preferences in using hand or foot are associated with other brain asymmetries such as emotional perception, language organization and visuospatial skills [12-14]. Determining laterality is beneficial both for research and clinical purposes (e.g., development of laterality, rehabilitation, sports training, etc.).

Several studies have investigated footedness and its relation to gait and postural control. In a review by Sadeghi and colleagues [15], the authors indicated that in tasks that are bilateral (e.g., gait) one foot would perform a mobilizing role (dominant), while the other foot would perform a stabilizing role (non-dominant). Further, the authors reported that many studies have found asymmetries in gait between the left and right legs among healthy non-disabled individuals. These differences were assessed via spatio-temporal, kinematic, kinetic and electromyography (EMG) parameters; and included differences in muscle strength and forces in both sagittal and frontal planes and differences in EMG amplitudes [15]. Additionally, it has been reported that there are anatomical differences between the lower limbs. In a study by Tate et al. [16], the authors reported that among young athletes the dominant leg had a larger vastus medialis muscle, while the non-dominant leg had a larger vastus lateralis muscle. Furthermore, in a comprehensive study by Ingelmark [17], the author reported that among individuals aged 6-13 years, 85% of right-handed subjects had a longer right leg and among those aged 14-20 years who are also right-handed they had a longer left leg. Also, the author added that for left-handed individuals they showed the opposite, with a longer left leg for the 6-13 years group, and a longer right leg for the 14-20 age group [17].

Evaluating footedness is of great significance for rehabilitation. Evidence supports that leg preference can influence postural control [18]. Indeed, as mentioned above, previous studies have reported asymmetries between the left and right leg during gait [15].

Rehabilitation specialists commonly use one lower extremity (e.g., the non-hemiparetic lower extremity among people post-stroke) as a reference point for therapy. As such, it is important to recognise the structural and functional differences between the dominant and non-dominant limbs. For people post-stroke, footedness shows a relationship with the severity of stroke-related impairments [19]. Among individuals with mild to moderate hemiparesis, the lower limb that is primarily used for support during an upright stance is determined by preference, while among those with more severe impairments, the supporting lower extremity is determined by convenience [19]. Thus, footedness among people post-stroke is determined via severity and should be assessed for optimal rehabilitation outcomes.

Many studies have investigated footedness and the findings of these studies indicate that lateral preference should be assessed using multi-item inventories and not use a single-item question (e.g., writing for handedness) [1-11]. The Waterloo Footedness Questionnaire-Revised (WFQ-R), is a multi-item, self-reported foot preference questionnaire originally developed in English by Elias and colleagues [20]. The WFQ-R is one of the most commonly used tools to determine foot preference. It is easy to administer and interpret; and has been previously used with a variety of populations, including non-disabled healthy adults [21], older adults [22], and individuals with neurological impairments [23]. There are 13 questions in the WFQ-R, which are answered on a 5-level Likert-type scale to determine which foot is the most often used. Responses are assigned a value between -2 and 2, with scores closer to 0 reflect equal foot preference, score closer to -2 indicate left foot preference, and 2 right foot preference. The total score of the questionnaire is used to categorize respondents as left-footed (score of -7 or less), mixed-footed (score of -6 to +6), or right-footed (score of +7 or higher) [21].

The WFQ-R has not been translated and cross-culturally adapted to the Arabic population. Given the importance of assessing footedness, the purpose of the current study is to translate the WFQ-R to the Arabic language, adapt it to the Saudi culture and investigate its validity and reliability.

## METHODS

### Study design

This is a longitudinal study that aimed to cross-culturally adapt the WFQ-R into Arabic. This study was conducted in accordance with the guidelines put forth by Beaton and colleagues [24]. The five stages suggested by the authors are: (i) translation of the questionnaire by two independent translators, (ii) synthesizing the translations, (iii) back translation to English by two native English speakers, (iv) expert committee review; and (v) pre-testing the translated questionnaire to evaluate comprehension. Afterwards, the final Arabic-version of the WFQ-R was tested in a large scale to establish its measurement properties.

Prior to the commencement of our study, one of the researchers (Dr. Lorin Elias) who developed the WFQ-R was contacted to obtain permission to translate the questionnaire. The local Research Ethics Committee approved all procedures. All participants provided an informed consent.

### Participants and recruitment

Using convenient sampling approach, participants recruitment was carried out in the community via advertisement posters and word of mouth. A total of 320 participants (30 participated in pilot testing) were included if they were adults (≥ 18 years of age), able to read, speak, and understand Arabic. Those who were unable to read or speak Arabic were excluded from this study. All participants were recruited from all regions in Saudi Arabia.

### Procedures

The translation and cross-cultural adaptation of the WFQ-R into Arabic was conducted in accordance with the guidelines put forth by Beaton and colleagues. Two independent forward translations were performed by two native Arabic speakers. The two produced forward translated versions were then synthesized into one synthesized version.

The synthesized version was back translated into English by two independent bilingual translators. An expert committee: including two methodologists and two language professionals and all forward and backward translators, was formed to review the translation and adaptation process. The expert committee suggested minor linguistic and idiomatic changes and have reached consensus that the reproduced back translated English version of the culturally adapted WFQ-R was compatible with the original one. After that, the pre-final version of the Arabic translated WFQ-R was created and was ready to be field-tested with participants.

A pilot study was conducted on the pre-final Arabic version of the WFQ-R on 30 participant who follow the recruitment criteria. Participants were asked to independently complete the Pre-final Arabic version of the WFQ-R. They were asked to respond freely and honestly to all items in the questionnaire and then were interviewed independently. After the 30-participant pilot testing, the final Arabic version of the revised Waterloo Footedness Questionnaire (WFQ-Ar) was reached.

The WFT-Ar was then tested further to evaluate its measurement properties including internal consistency, reliability, and validity. Two-hundred and ninety Arabic speakers were asked to complete a general information sheet for demographic data after consenting to participate in this study. This information sheet had a commonly used question about foot dominancy stated as “which foot would you choose to kick this ball in front of you?” with answering options of “right” or left”. After that, participants were asked to fill the WFQ-Ar twice (one week apart).

### Data analyses

All data collected were analysed for exploring missing data and descriptive statistics including participants’ characteristics, foot dominancy based on the information sheet question, footedness based on the WFQ-Ar. Face validity was determined upon participants’ responses to the interview questioning on relevance and appropriateness of the scale to determine foot dominancy. The content validity was established if the expert committee members reached a consensus concerning the relevance and appropriateness of the scale to determine the footedness of Arabic speakers. The floor and ceiling effects of the WFQ-Ar were determined by computing the percentage of participants scoring lowest or highest. The scale was considered to have flooring or ceiling effect when more than or equal to 15% of the participants had the lowest or highest possible score.

Construct validity of the WFQ-Ar was determined using Factor analysis with direct oblimin rotation method to extract factors of eigenvalues greater than Kaiser’s criterion of one [25].

Cronbach’s α was used to evaluate the internal consistency of the WFQ-Ar, where a value of α = 0.7 was considered the minimum for an adequate consistency. While the interclass correlation coefficient for absolute agreement (ICC_2,1_) was used to assess test-retest reliability, where a minimum ICC value of 0.70 was considered an adequate test-retest reliability for the WFQ-Ar. The standard error of measurement (SEM) was used to examine the scale measurement error associated with the test-retest examination. The SEM was computed using the formula 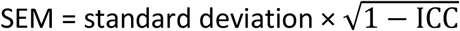.

Hypothesis testing was used for construct validation. The WFQ-Ar was hypothesized to have moderate to strong positive correlation (r ≥ 0.4) with the dominancy question answered by participants. The spearman rank coefficient (r_s_) was used to examine the hypothesized correlation for construct validity. All data analyses were calculated using IBM SPSS Statistics for windows, version 26 (IBM Corp. Armonk, NY) and were deemed significant at α < 0.05.

## RESULTS

Two hundred and ninety Arabic speakers participated in this study. The detailed participants’ characteristics are shown in Table 1. The translation and cross-cultural adaptation process of the WFQ-Ar was straight forward with the inclusion of 30 Arabic speaking participants and eight expert committee members. Only item number 9 “Which foot would you use to help push a shovel into the ground?” that needed to be expressed with its conceptual meaning by the addition of more words. Also, the resulted cultural adaptation changed the answering grid from using abbreviations (La/Ru) into actual words (Left always/Right usually) to make it more clear for the respondent. All 30 participants were interviewed after answering the pre-final version of the WFQ-Ar indicating that the questionnaire was clear, relevant, and appropriate to determine foot dominancy. Their testament supported the adequacy of the WFQ-Ar face validity. Furthermore, the expert committee have reached consensus concerning the relevance and appropriateness of the WFQ-Ar for Arabic speakers to determine foot dominancy. Also, the completeness of the WFQ-Ar items was satisfactory and the absence of floor and ceiling effects in the analysis further support adequate content validity.

**Table 1:** Participants’ characteristics; n=290

|  |  |
| --- | --- |
| **Age** in years (mean ± SD) | 29.9 ± 11.3 |
| **Gender** |  |
| Male | 133 (45.9%) |
| Female | 157 (54.1%) |
| **Education Status** |  |
| No Education | 1 (0.3%) |
| Elementary | 5 (1.7%) |
| Intermediate | 19 (6.6%) |
| High School | 103 (35.5%) |
| University Graduate | 150 (51.7%) |
| Higher Education | 12 (4.1%) |
| **Employment Status** |  |
| Student | 132 (45.5%) |
| Employed | 101 (34.8%) |
| Unemployed | 57 (19.7%) |
| **Dominant Foot\*** |  |
| Left | 28 (9.7%) |
| Right | 262 (90.3%) |

\* Dominant Foot based on answers to the question *"Which foot you use to kick a ball aiming for a goal in front of you?"*

For construct validity, The Kaiser-Meyer-Olkin measure of sampling adequacy was met (p = 0.93), Bartlett’s test of sphericity was satisfied (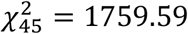, p < 0.001), and the determinant score suggests a lack of multicolinearity (r = 0.112). These three findings suggested that the data were appropriate for a factor analysis to be utilized. Exploratory factor analysis indicated that only one factor had an eigenvalue greater than Kaisar’s criterion of one (6.01) explaining 60.05% of the variance (Table 2). After extraction, the factor explained 55.79% of the variance. Thus, all items of the WFQ-Ar have one main dimension (Figure 1). Furthermore, all 10 items loaded more than 0.5 on that single factor with loadings ranging from 0.56 to 0.82 (Table 3).

**Table 2:** WFQ-Ar Factor Structure

| Factor | Initial Eigenvalue |  |  | After Extraction |  |  |
| --- | --- | --- | --- | --- | --- | --- |
|  | Total | % of Variance | Cumulative % | Total | % of Variance | Cumulative % |
| 1 | 6.01 | 60.79 | 60.05 | 5.58 | 55.79 | 55.79 |
| 2 | 0.82 | 8.18 | 68.23 |  |  |  |
| 3 | 0.63 | 6.27 | 74.49 |  |  |  |
| 4 | 0.50 | 5.04 | 79.53 |  |  |  |
| 5 | 0.47 | 4.65 | 84.18 |  |  |  |
| 6 | 0.41 | 4.13 | 88.31 |  |  |  |
| 7 | 0.38 | 3.75 | 92.06 |  |  |  |
| 8 | 0.29 | 2.92 | 94.98 |  |  |  |
| 9 | 0.28 | 2.78 | 97.75 |  |  |  |
| 10 | 0.26 | 2.25 | 100.00 |  |  |  |

**Figure 1:**
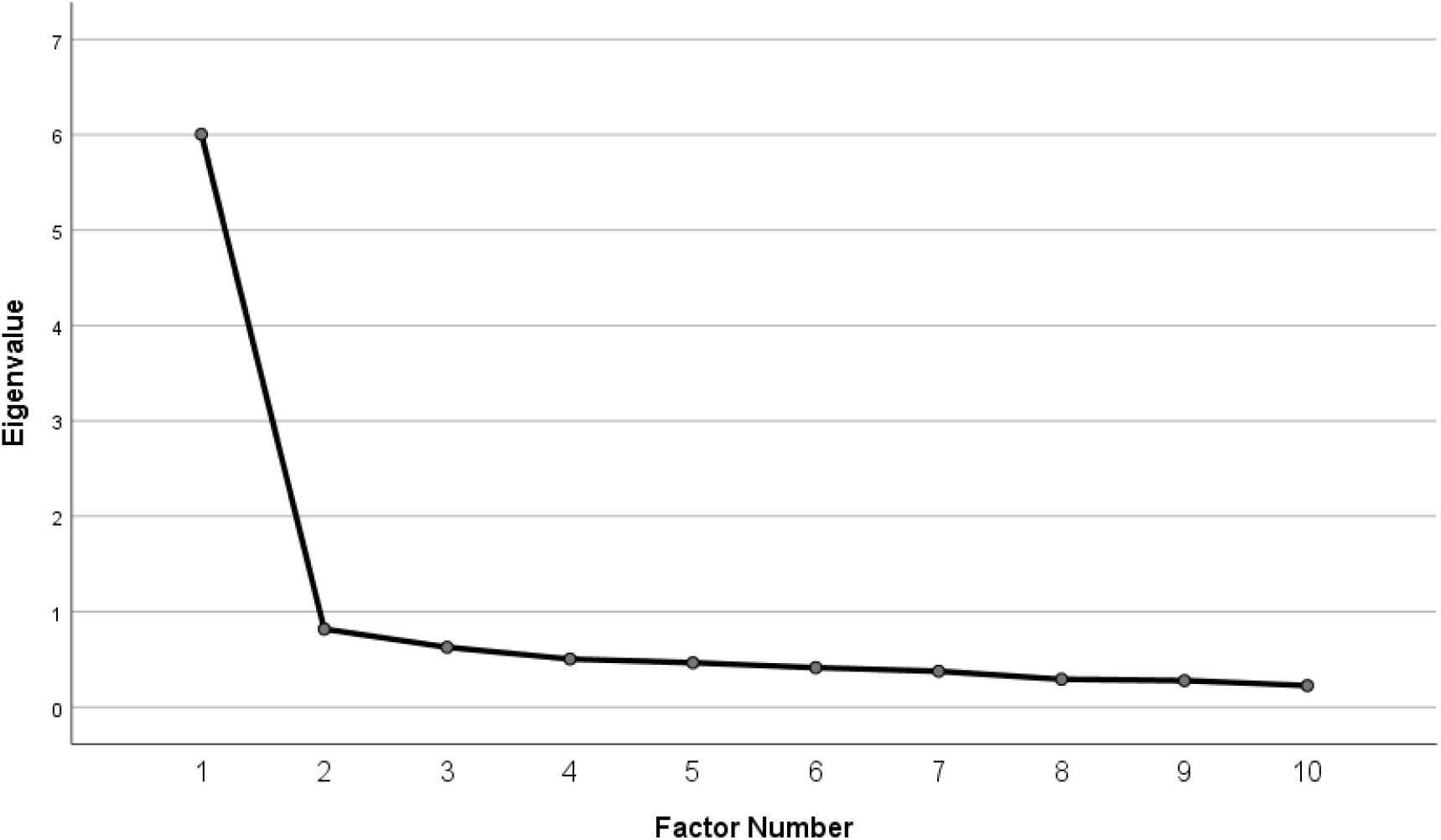
Exploratory factor analysis figure showing the factor number on the horizontal axis and the eigenvalue on the vertical axis.

**Table 3:** Internal consistency, rotated factor loadings for the WFQ-Ar 10 items using principal axis method with direct oblimin rotation, and the explained variance for the factor, N = 290

| Questionnaire<br>(Single Factor) | Cronbach's $\alpha$ | Items | Cronbach's $\alpha$<br>if item<br>deleted | Rotated<br>Factor<br>Loadings |
| --- | --- | --- | --- | --- |
| Footedness | 0.932 |  |  |  |
|  |  | Item 1 | 0.924 | 0.82 |
|  |  | Item 2 | 0.926 | 0.82 |
|  |  | Item 3 | 0.924 | 0.78 |
|  |  | Item 4 | 0.925 | 0.77 |
|  |  | Item 5 | 0.923 | 0.77 |
|  |  | Item 6 | 0.922 | 0.74 |
|  |  | Item 7 | 0.923 | 0.73 |
|  |  | Item 8 | 0.926 | 0.73 |
|  |  | Item 9 | 0.925 | 0.71 |
|  |  | Item 10 | 0.927 | 0.56 |

The internal consistency of the WFQ-Ar was excellent with Cronbach’s alpha of 0.93. With regards to WFQ-Ar stability, results showed excellent test-retest reliability with ICC value of 0.94 (95% CI=0.93-0.95). The Bland-Altman plot showed acceptable agreement between test and retest scores (Figure 2). Also, the scale measurement error computed from the test-retest reliability (SEM) is of 4.5 points. Further, the hypothesized correlation between the WFQ-Ar and the participant’s dominancy answering was found to be significant with r = 0.48, p < 0.001.

**Figure 2:**
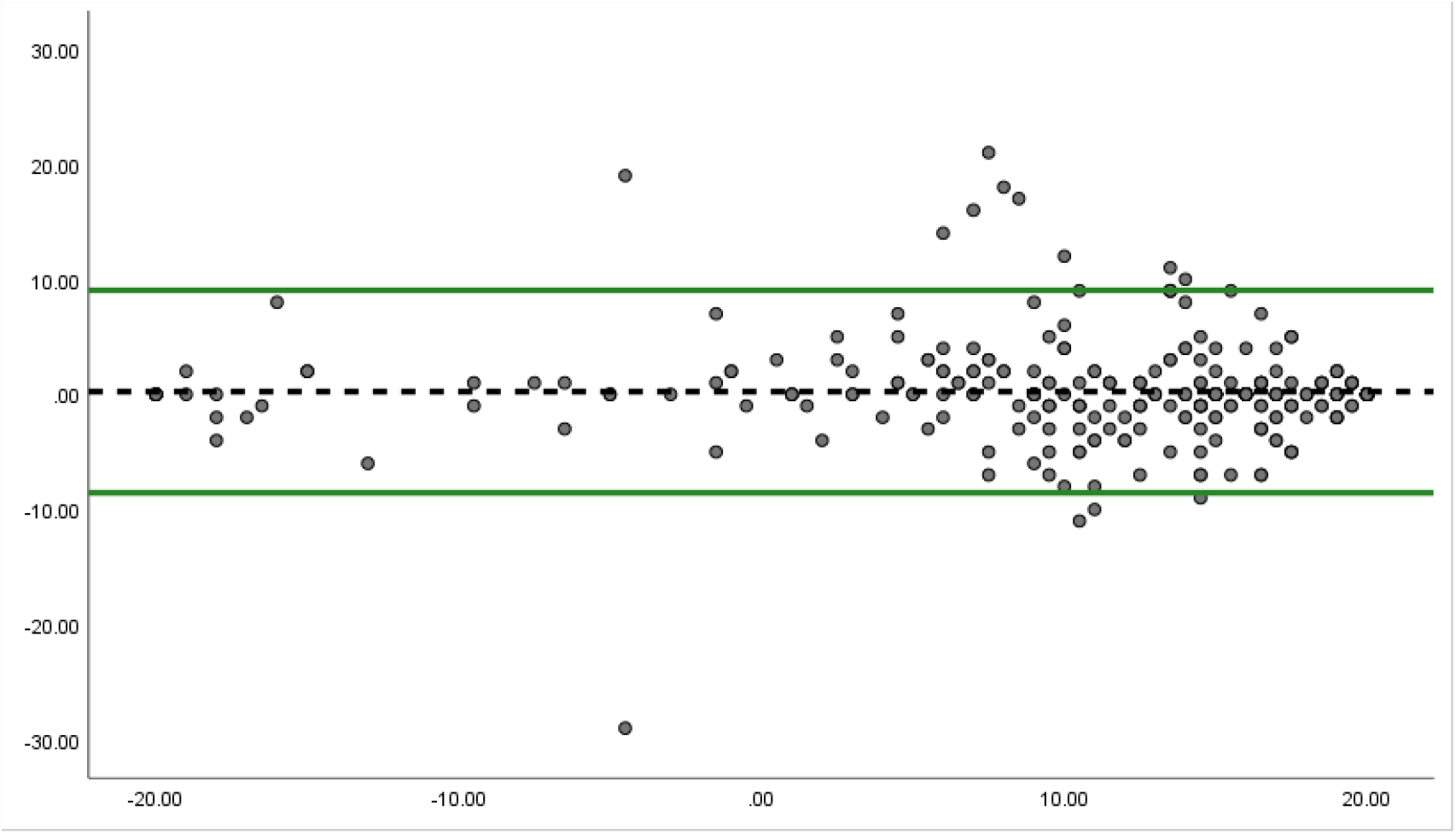
Bland-Altman plot.

## DISCUSSION

The current study was conducted to culturally adapt the WFQ-R into the Arabic language and to test its measurement properties including validity, internal consistency, test-retest reliability and measurement error. It was hypothesized that the WFQ-Ar would have good internal consistency, excellent test-retest reliability, and would be a valid measure of footedness. The result of this study supports these hypotheses.

The cross-cultural adaptation process of the WFQ-R was smooth, with only one change to the wording of one item in the questionnaire and omitting the abbreviations for the answering grid. This change was agreed upon by the expert committee and was done to improve the clarity of the WFQ-R for Arabic speakers.

The results of cross-cultural adaptation appear to support the face validity of the WFQ-Ar. The expert committee involved in translating the WFQ-R reached consensus that the WFQ-Ar is both relevant and appropriate for assessing footedness. Further, face validity was also supported by the testaments of participants who all indicated that the WFQ-Ar was clear and relevant and suitable for assessing footedness. For content validity, the results of the analyses indicate that the WFQ-Ar has no floor or ceiling effect, indicating adequate content validity [26]. The concurrent validity was also tested as the WFQ-Ar was significantly correlated with the criterion “which foot would you choose to kick this ball in front of you?”. Suggesting that the WFQ-Ar can predict footedness similar to how clinicians commonly assess foot preference.

Factor analysis was used to examine the construct validity of the WFQ-Ar, and it was anticipated that the WFQ-Ar would have one factor structure. The results of the factor analysis confirmed that the WFQ-Ar has one major factor underlying its items. All items within the WFQ-Ar loaded significantly on this factor. As such, there was a great proportion of the total variance (55.79%) of the WFQ-Ar. The overall results of the factor analysis verify the construct validity of the WFQ-Ar and support that this measure primarily assesses one underlying construct; footedness. The results related to construct validity found in the current study are similar to what is previously reported [27].

The Arabic version of the WFQ-R showed excellent internal consistency (Cronbach’s alpha=0.93) indicating that items within the scale are correlated, homogenous and are not redundant [28]. Cronbach’s alpha of the WFQ-Ar did not change with deletion of items one at a time (Table 3). The consistency of Cronbach’s alpha indicate that all items are homogenous and correlated well with each other and that removal of any item would not improve the internal consistency and homogeneity of the scale.

The reliability analysis confirmed that the WFQ-Ar is reliable with excellent test-retest reliability (ICC=0.94). The magnitude of reliability found in the current study is higher than that reported for the Brazilian (Portuguese) version, and the Chinese version. The Standard error of measurement (SEM) of the WFQ-Ar was 4.5 points. Expressed as a percentage of the overall WFQ-Ar score, SEM was 10%. The SEM, based on its value and percentage relative to overall score, the measurement error of WFQ-Ar presented in this study seems to be clinically applicable. Lastly, the Bland-Altman plot showed adequate agreement between test and retest scores, with over 95% of the scores between the agreement lines; indicating excellent reliability of the WFQ-Ar.

The cross-cultural adaptation and investigation of measurement properties of the WFQ-Ar performed in this study provides both clinicians and researchers in the Arabic-speaking countries with a validated tool that can be used to assess footedness. As mentioned above, clinicians often need to assess footedness in order to establish a reference point for therapeutic programs. Unfortunately, clinicians commonly assess footedness using a single-item question. However, lateral preference should be assessed via multi-item questionnaires as lateral preference is a spectrum rather than dichotomous [1-11].

In the current study the WFQ-R was translated into modern standard Arabic language and did not utilize any local dialects. The translation was performed in this manner to encourage its use throughout the Arabic-speaking countries, as modern standard Arabic language is understood across these countries. However, it should be mentioned that all participants recruited in the current study were Saudi Arabian. Therefore, using the WFQ-Ar in other Arabic-speaking countries should be preceded by formal testing of its measurement properties in the local context. Moreover, the WFQ-Ar was tested among adult, non-disabled individuals. Further investigation of the WFQ-Ar measurement properties should be performed among other populations.

## CONCLUSION

In conclusion, this study was carried out to culturally adapt the WFQ-R and to examine measurement properties of the Arabic version of this questionnaire. The adaptation process was smooth with slight modification to the original WFQ-R. Participants reported that the WFQ-Ar was clearly understood and easy to fill. The WFQ-Ar is a valid and reliable measure for determining footedness. These measurement properties of the WFQ-Ar affirm the clinical usefulness of this measure for clinical and research purposes.

## Data Availability

All data produced in the present work are contained in the manuscript

## Acknowledgement

The author would like to acknowledge Ms. Hanan Aloraini for her important contribution to this manuscript.

## REFERENCES

1. Dittmar M. Functional and postural lateral preferences in humans: interrelations and life-span age differences. Hum Biol. 2002;74(4):569–85. doi: 10.1353/hub.2002.0040.

2. Bourassa DC, McManus IC, Bryden MP. Handedness and eye-dominance: a meta-analysis of their relationship. Laterality. 1996;1(1):5–34. doi: 10.1080/713754206.

3. Dellatolas G, Curt F, Dargent-Pare C, De Agostini M. Eye dominance in children: a longitudinal study. Behav Genet. 1998;28(3):187–95. doi: 10.1023/a:1021471129962.

4. Kang Y, Harris LJ. Handedness and footedness in Korean college students. Brain Cogn. 2000;43(1-3):268–74.

5. McManus IC, Porac C, Bryden MP, Boucher R. Eye-dominance, writing hand, and throwing hand. Laterality. 1999;4(2):173–92. doi: 10.1080/713754334.

6. Noonan M, Axelrod S. Earedness (ear choice in monaural tasks): its measurement and relationship to other lateral preferences. J Aud Res. 1981;21(4):263–77.

7. Porac C. Eye preference patterns among left-handed adults. Laterality. 1997;2(3-4):305–16. doi: 10.1080/713754270.

8. Reiss M. [Current aspects of ear preference]. Folia Phoniatr Logop. 1998;50(1):19–27. doi: 10.1159/000021446.

9. Reiss M, Reiss G. Earedness and handedness: distribution in a German sample with some family data. Cortex. 1999;35(3):403–12. doi: 10.1016/s0010-9452(08)70808-6.

10. Suar D, Mandal MK, Misra I, Suman S. Lifespan trends of side bias in India. Laterality. 2007;12(4):302–20. doi: 10.1080/13576500701282630.

11. Tran US, Stieger S, Voracek M. Evidence for general right-, mixed-, and left-sidedness in self-reported handedness, footedness, eyedness, and earedness, and a primacy of footedness in a large-sample latent variable analysis. Neuropsychologia. 2014;62:220–32. doi: 10.1016/j.neuropsychologia.2014.07.027.

12. Watson GS, Pusakulich RL, Ward JP, Hermann B. Handedness, footedness, and language laterality: evidence from Wada testing. Laterality. 1998;3(4):323–30. doi: 10.1080/713754311.

13. Whitehouse AJ, Bishop DV. Hemispheric division of function is the result of independent probabilistic biases. Neuropsychologia. 2009;47(8-9):1938–43. doi: 10.1016/j.neuropsychologia.2009.03.005.

14. Haddad JM, Rietdyk S, Ryu JH, Seaman JM, Silver TA, Kalish JA, et al. Postural asymmetries in response to holding evenly and unevenly distributed loads during self-selected stance. J Mot Behav. 2011;43(4):345–55. doi: 10.1080/00222895.2011.596169.

15. Sadeghi H, Allard P, Prince F, Labelle H. Symmetry and limb dominance in able-bodied gait: a review. Gait & posture. 2000;12(1):34–45. doi: 10.1016/s0966-6362(00)00070-9.

16. Tate CM, Williams GN, Barrance PJ, Buchanan TS. Lower extremity muscle morphology in young athletes: an MRI-based analysis. Med Sci Sports Exerc. 2006;38(1):122–8. doi: 10.1249/01.mss.0000179400.67734.01.

17. Ingelmark B. Asymmetries in the length of the extremities and their relation to right-and left-handedness. Upsala Laekarefoerening Forhandlingar. 1974;52:17–82.

18. Emery CA, Cassidy JD, Klassen TP, Rosychuk RJ, Rowe BB. Development of a clinical static and dynamic standing balance measurement tool appropriate for use in adolescents. Phys Ther. 2005;85(6):502–14.

19. Mundim AC, Paz CC, Fachin-Martins E. Could be the predominantly-used hemibody related to the weight bearing distribution modified by the chronic hemiparesis after stroke? Med Hypotheses. 2015;85(5):645–9. doi: 10.1016/j.mehy.2015.08.007.

20. Elias LJ, Bryden MP, Bulman-Fleming MB. Footedness is a better predictor than is handedness of emotional lateralization. Neuropsychologia. 1998;36(1):37–43.

21. Grouios G, Hatzitaki V, Kollias N, Koidou I. Investigating the stabilising and mobilising features of footedness. Laterality. 2009;14(4):362–80. doi: 10.1080/13576500802434965.

22. Aloraini SM, Glazebrook CM, Sibley KM, Singer J, Passmore S. Anticipatory postural adjustments during a Fitts’ task: Comparing young versus older adults and the effects of different foci of attention. Hum Mov Sci. 2019;64:366–77. doi: 10.1016/j.humov.2019.02.019.

23. Camargos MB, Palmeira ADS, Fachin-Martins E. Cross-cultural adaptation to Brazilian Portuguese of the Waterloo Footedness Questionnaire-Revised: WFQ-R-Brazil. Arq Neuropsiquiatr. 2017;75(10):727–35. doi: 10.1590/0004-282X20170139.

24. Beaton DE, Bombardier C, Guillemin F, Ferraz MB. Guidelines for the process of cross-cultural adaptation of self-report measures. Spine (Phila Pa 1976). 2000;25(24):3186–91. doi: 10.1097/00007632-200012150-00014.

25. Kaiser HF. The application of electronic computers to factor analysis. Educational and psychological measurement. 1960;20(1):141–51.

26. Terwee CB, Bot SD, de Boer MR, van der Windt DA, Knol DL, Dekker J, et al. Quality criteria were proposed for measurement properties of health status questionnaires. J Clin Epidemiol. 2007;60(1):34–42. doi: 10.1016/j.jclinepi.2006.03.012.

27. Kapreli E, Athanasopoulos S, Stavridis I, Billis E, Strimpakos N. Waterloo Footedness Questionnaire (WFQ-R): cross-cultural adaptation and psychometric properties of Greek version. Physiotherapy. 2015;101:e721.

28. De Vet HC, Terwee CB, Mokkink LB, Knol DL. Measurement in medicine: a practical guide: Cambridge University Press; 2011.

